# Predictors of Hepatitis B Vaccine Uptake among Healthcare Workers in Sokoto, Nigeria

**DOI:** 10.1101/2025.06.22.25330077

**Authors:** Suleiman Idris Ahmad, Khadeejah Liman Hamza, Yahya Mohammed, Sa’adatu Shinkafi, Abubakar Maiyaki, Nuhu Aliyu, Muhammad Shakir Balogun, Abdulhakeem Olorukooba, Chukwuma Umeokonkwo, Mustapha Umar Imam, Lubabatu Abdulazeez, Habila Yunana, Lawal Aminu, Kabir Sabitu

## Abstract

**Background:** Hepatitis B virus (HBV) infection is a major occupational hazard for healthcare workers (HCWs) in Nigeria, a high-prevalence country. With the HBV vaccine only integrated into the national childhood immunization program in 2004, most current HCWs are potentially unvaccinated and unprotected. This study aimed to determine the hepatitis B vaccine uptake rate and identify its predictors among HCWs in Sokoto State, Nigeria.

**Methods:** An analytical cross-sectional study was conducted between January and March 2023. A total of 804 HCWs were selected from primary, secondary, and tertiary health facilities using a multistage sampling technique. Data on sociodemographic characteristics, knowledge, attitude, and vaccine uptake were collected via a structured questionnaire. Multivariate logistic regression was used to identify independent predictors of vaccine uptake.

**Results:** Only 35.5% (285/804) of HCWs reported receiving at least one dose of the hepatitis B vaccine. Of these, just 51.6% (147/285) were fully vaccinated with three or more doses. Significant positive predictors of uptake included having a positive attitude towards the vaccine (aOR: 1.84; 95% CI: 1.19-2.84) and an employer recommending the vaccine at employment (aOR: 1.85; 95% CI: 1.15-2.98). Negative predictors included being a health assistant (aOR: 0.15; 95% CI: 0.03-0.91), working in a tertiary facility (aOR: 0.47; 95% CI: 0.24-0.93), having poor knowledge of HBV (aOR: 0.22; 95% CI: 0.07-0.69), and the absence of institutional advocacy (aOR: 0.62; 95% CI: 0.30-0.72).

**Conclusion:** Hepatitis B vaccination coverage among HCWs in Sokoto is critically low, exposing them to significant occupational risk. There is an urgent need for health authorities and hospital management to implement mandatory vaccination policies, coupled with targeted educational programs for vulnerable cadres, to improve uptake and protect this vital workforce.

## Introduction

Hepatitis B virus (HBV) infection is a significant global public health challenge. In 2015, the World Health Organisation (WHO) estimated that 257 million people were living with chronic HBV infection, which led to approximately 887,000 deaths, primarily from complications such as liver cirrhosis and hepatocellular carcinoma^2^. The WHO African Region has one of the highest prevalences of HBV, with an estimated 6.1% of the adult population infected^2^. In Nigeria, the burden is particularly high; a 2016 national survey reported an HBV prevalence of 12.2%, while the more recent Nigeria HIV/AIDS Indicator and Impact Survey (NAIIS) in 2020 found a prevalence of 8.1%^3,^□. HBV, a DNA virus from the hepadnaviridae family, is known to cause both acute and chronic hepatitis, which can progress to severe liver disease^1^.

Healthcare workers (HCWs) represent a high-risk group for contracting HBV due to the nature of their work. Their clinical environment and frequent contact with patients’ blood and other potentially infectious body fluids make them particularly vulnerable to blood-borne pathogens□,□. HBV is highly infectious—more so than HIV—and can remain viable on environmental surfaces for at least seven days, posing a persistent threat in clinical settings where environmental cleaning may be suboptimal□. Globally, it is estimated that thirty-seven per cent of HBV infections are attributable to occupational exposure.

Immunization remains the single most effective intervention for preventing HBV infection in susceptible individuals. The hepatitis B vaccine is safe and highly effective, offering 98% to 100% protection against the development of chronic infection and its complications^11,12^. Despite its availability, a critical gap in protection exists for a large segment of the Nigerian healthcare workforce. The HBV vaccine was not integrated into Nigeria’s National Programme on Immunization (NPI) for infants until 2004. Consequently, most HCWs currently in practice, being older than 18 years, would have missed the opportunity for vaccination during childhood. This has created a large, potentially susceptible adult population at high occupational risk. Compounding this issue is the lack of a formal national policy for the screening and mandatory vaccination of HCWs in Nigeria□.

Given the low historical immunization coverage in Northern Nigeria and the unique vulnerability of the current healthcare workforce, understanding the factors that influence vaccine uptake is critical for developing effective public health strategies^1^□. Therefore, this study aimed to determine the hepatitis B vaccine uptake rate and identify the sociodemographic, institutional, and personal factors predicting its uptake among healthcare workers in Sokoto State, Nigeria.

## Methods

### Study Design and Setting

This analytical cross-sectional study was conducted between January 16 and March 14, 2023, among healthcare workers (HCWs) in Sokoto State, located in Northwestern Nigeria. The state capital, Sokoto City, hosts a mix of public and private health facilities, including tertiary, secondary, and primary levels of care. At the time of the study, there was no formal vaccination program for HCWs in Sokoto State.

### Study Population and Sampling

The study population comprised approximately 6,521 HCWs distributed across 86 health facilities within Sokoto City. The inclusion criteria encompassed all clinical healthcare workers (such as doctors, nurses, laboratory staff, and health officers) and health assistants working in both public and private facilities. Pharmacists and individuals who were unavailable during the study period were excluded.

The sample size was calculated using the Cochran formula□^1^, based on a hepatitis B vaccine uptake prevalence of 40.3% from a previous study in Sokoto^3^□, a 99% confidence level, a 3.5% margin of error, and a 10% non-response rate, yielding a target sample size of 893 participants.

A multistage sampling technique was employed. In the first stage, health facilities were stratified by tier, and a proportionate number were randomly selected: ten primary, five secondary, and two tertiary facilities. In the second stage, HCWs within the selected facilities were chosen by balloting, with proportionate allocation based on the staff population of each facility. Of the 893 HCWs approached, 804 completed the questionnaire, resulting in a response rate of 90%.

### Data Collection and Instrument

Data were collected using a standardized, pre-tested, and validated semi-structured questionnaire, which was administered by trained interviewers. The questionnaire gathered information on sociodemographic characteristics, knowledge and awareness of HBV, risk perception, attitudes towards the virus and vaccine, occupational exposure to HBV, and existing advocacy and employment policies at the health facilities. Data were collected electronically, and unique codes were assigned to each participant to ensure anonymity.

### Variables and Measurement

The primary outcome variable was self-reported hepatitis B vaccine uptake, defined as having received at least one dose of the vaccine (’Yes’ or ’No’). Key independent variables included sociodemographic characteristics. Composite scores were created for knowledge, attitude, and risk perception. Knowledge was assessed using 15 questions, with scores categorized as ’Good’ (11-15), ’Fair’ (6-10), or ’Poor’ (0-5). Attitude and risk perception were measured using a 5-point Likert scale and categorized as ’Positive’ or ’Negative’ based on score distributions.

### Statistical Analysis

Data were cleaned and analyzed using Epi Info. Descriptive statistics, including frequencies and proportions, were calculated to summarize the variables. Bivariate analysis using the Chi-square test was conducted to assess associations between independent variables and vaccine uptake. Subsequently, multivariate binary logistic regression was performed to identify independent predictors of vaccine uptake. Adjusted odds ratios (aOR) with their corresponding 95% confidence intervals (CI) were computed, and a p-value of <0.05 was considered statistically significant.

### Ethical Considerations

The study was conducted in accordance with ethical standards, receiving clearance from the Sokoto State Ministry of Health (Ref: SMH/1580/V.IV) and the Usmanu Danfodiyo University Teaching Hospital, Sokoto (Ref: NHREC/30/012/2019). Permission was also obtained from the management of the selected health facilities. All participants provided written informed consent before the interviews. Participation was voluntary, and confidentiality was maintained by de-identifying all data.

## Results

### Sociodemographic Characteristics of Participants

A total of 804 healthcare workers (HCWs) participated in the study. The median age of the respondents was 35 years (Interquartile Range: 28-43). A proportion of participants were female (54.6%, n=439), and most were married (66.4%, n=534). The most represented professional cadres were nurses and midwives (34.1%, n=274), followed by laboratory staff (24.5%, n=197) and health assistants (21.5%, n=173). A significant proportion of the HCWs worked in tertiary health facilities (62.7%, n=504), and the median duration of professional practice was 6 years (Interquartile Range: 2-13). Table 1 summarizes the sociodemographic characteristics of the study population.

**Table 1:** Sociodemographic Characteristics of Healthcare Workers in Sokoto State (n=804)

| Characteristic | Category | Frequency (n) | Percentage (%) |
| --- | --- | --- | --- |
| **Sex** | Female | 439 | 54.6 |
|  | Male | 365 | 45.4 |
| **Age Group (years)** | 17 - 26 | 159 | 19.8 |
|  | 27 - 36 | 312 | 38.8 |
|  | 37 - 46 | 193 | 24.0 |
|  | ≥ 47 | 140 | 17.4 |
| **Professional Cadre** | Doctors and Dentists | 76 | 9.5 |
|  | Nurses and Midwives | 274 | 34.1 |
|  | Laboratory Staff | 197 | 24.5 |
|  | Health Officers | 84 | 10.4 |
|  | Health Assistants | 173 | 21.5 |
| **Tier of Health Facility** | Primary | 106 | 13.2 |
|  | Secondary | 194 | 24.1 |
|  | Tertiary | 504 | 62.7 |
| **Highest Education** | No formal / Primary / Secondary | 173 | 21.5 |
|  | Tertiary (Diploma, Bachelors, etc.) | 631 | 78.5 |

### Hepatitis B Vaccine Uptake

The self-reported uptake of at least one dose of the hepatitis B vaccine was 35.5% (n=285). Among the 285 HCWs who had initiated vaccination, only 51.6% (n=147) reported having received three or more doses, constituting full vaccination. The primary motivation for vaccination among those who received it was a fear of contracting hepatitis B (77.2%). For the 519 unvaccinated participants, the most cited obstacle was not knowing where to get the vaccine (33.1%).

### Predictors of Hepatitis B Vaccine Uptake

Table 2 presents the results of the multivariate logistic regression analysis, which identified several independent predictors of hepatitis B vaccine uptake. Factors that positively predicted vaccine uptake included being in the age group of 27-36 years (aOR: 2.33; 95% CI: 1.06-5.10) or 37-46 years (aOR: 2.44; 95% CI: 1.23-4.85) when compared to those aged 47 and older. HCWs who had a positive attitude towards the vaccine and its importance were more likely to be vaccinated (aOR: 1.84; 95% CI: 1.19-2.84). Furthermore, an employer recommending the need for the hepatitis B vaccine at the time of employment significantly increased the likelihood of uptake (aOR: 1.85; 95% CI: 1.15-2.98).

**Table 2:** Multivariate Logistic Regression of Predictors of Hepatitis B Vaccine Uptake.

| **Characteristic** | **Category** | **aOR (95% CI)** | **P-value** |
| --- | --- | --- | --- |
| **Age Group (years)** | 27 - 36 | 2.33 (1.06 - 5.10) | 0.035 |
|  | 37 - 46 | 2.44 (1.23 - 4.85) | 0.011 |
| | $\geq 47$ | *Reference* | |
| **Relationship Status** | Single vs. Married | 0.51 (0.29 - 0.87) | 0.014 |
| **Professional Cadre** | Health Assistant vs. Doctor/Dentist | 0.15 (0.03 - 0.91) | 0.039 |
| **Tier of Health Facility** | Tertiary vs. Primary | 0.47 (0.24 - 0.93) | 0.030 |
| **Knowledge Score** | Poor vs. Fair | 0.22 (0.07 - 0.69) | 0.010 |
| **Attitude Score** | Positive vs. Negative | 1.84 (1.19 - 2.84) | 0.006 |
| **Institutional Advocacy** | Absence vs. 'Don't Know' | 0.62 (0.30 - 0.72) | 0.001 |
| **Employer Policy** | Recommended Vaccine vs. Not | 1.85 (1.15 - 2.98) | 0.012 |

*aOR = Adjusted Odds Ratio; CI = Confidence Interval.*

Conversely, several factors were associated with lower odds of vaccination. HCWs who were single were less likely to be vaccinated than those who were married (aOR: 0.51; 95% CI: 0.29-0.87). When compared to doctors and dentists, health assistants were significantly less likely to have received the vaccine (aOR: 0.15; 95% CI: 0.03-0.91). Working in a tertiary health facility was associated with a lower likelihood of vaccination compared to working in a primary facility (aOR: 0.47; 95% CI: 0.24-0.93). Lastly, having poor knowledge about HBV (aOR: 0.22; 95% CI: 0.07-0.69) and working in an institution with no advocacy measures for vaccination (aOR: 0.62; 95% CI: 0.30-0.72) were also significant negative predictors.

## Discussion

This study identified a critically low uptake of the hepatitis B vaccine among healthcare workers in Sokoto State, Nigeria, and uncovered key predictors influencing this trend. The findings indicate that despite a high-risk environment, a significant portion of the healthcare workforce remains unprotected against a major occupational hazard. The determinants of vaccine uptake were multi-faceted, involving a combination of individual factors like age and attitude, professional roles, and crucial institutional factors such as employer recommendations and the type of health facility.

The self-reported vaccine uptake rate of 35.5% in our study is alarmingly low and underscores a significant public health issue. This rate is lower than that reported in several other African studies^23,^□□,□□. When compared with other research within Nigeria, our finding is lower than the 40.3% reported in a 2015 study in Sokoto and the 43.4% found in Calabar, yet higher than the 14.2% uptake documented in a study in Enugu^22,3^□,□□. This suggests that while low HBV vaccination coverage among HCWs is a widespread national problem, local factors may influence the exact rates. Furthermore, this uptake is markedly lower than the coverage seen in studies from Asia, Europe, and the Middle East highlighting a substantial gap in occupational health and safety practices in our setting^21,^□^1^,□^2^,□□.

Our analysis identified that HCWs aged between 27 and 46 years were more than twice as likely to be vaccinated. This finding is consistent with studies from Ghana and Enugu, which also showed higher uptake in more established career-age groups^□□,□□^. This may be attributable to increased awareness of occupational risks over time.

As expected, a positive attitude towards the vaccine was a strong predictor of uptake, emphasizing that HCWs’ beliefs and perceptions are crucial targets for behavioral interventions.

The most actionable finding is the powerful role of institutional prompts; an employer’s recommendation to vaccinate at the point of employment increased uptake by 85%. This finding, like a report from Pakistan, suggests that simple institutional cues to action may be more effective in driving behavior than relying on an individual’s internal motivation, such as fear of infection□^2^.

Conversely, several factors were associated with a lower likelihood of vaccination. Health assistants were 85% less likely to be vaccinated compared to doctors, pointing to a significant disparity in occupational health access and knowledge between different professional cadres. This finding contrasts with a Botswanan study that reported increased uptake in doctors and laboratory staff rather than health assistants□□.

A particularly interesting and counter-intuitive finding was that working in a tertiary facility was a negative predictor of vaccine uptake. This may be due to factors such as higher workload and less time for personal health maintenance, a hypothesis supported by an Enugu study where ’lack of time’ was a major reason for non-vaccination□□.

Finally, poor knowledge and the absence of institutional advocacy were also strong negative predictors, reinforcing that both individual awareness and a supportive workplace environment are essential for improving vaccination coverage.

The findings of this study have direct implications for policy and practice. The low overall uptake demonstrates a clear failure to protect a high-risk population and calls for urgent intervention. The strong predictive power of an employer’s recommendation provides compelling evidence that making hepatitis B vaccination a mandatory condition of employment for HCWs would be a highly effective strategy. Furthermore, the significant knowledge gaps and the specific vulnerability of cadres like health assistants highlight the need for targeted, continuous education programs that go beyond simple awareness to address specific barriers and misconceptions. Hospital management has a critical role to play in fostering a culture of safety, implementing clear vaccination policies, and removing obstacles to access.

This study has several strengths, including its large sample size, methodologically sound probability sampling, and the inclusion of a wide range of health facilities (public and private; primary, secondary, and tertiary), which enhances the generalizability of the findings within the study area. However, the study’s limitations must be considered. Its cross-sectional design means that causality cannot be established, and the reliance on self-reported vaccination status may be subject to recall bias. While the findings are robust for Sokoto state, their generalizability to all of Nigeria should be approached with caution.

## Conclusion

In conclusion, this study reveals that the uptake of the hepatitis B vaccine among healthcare workers in Sokoto, Nigeria, is critically inadequate, leaving a vast majority of this high-risk group unprotected. Vaccine uptake is not driven by a single factor but is a complex issue influenced by a combination of individual, professional, and institutional determinants. Positive predictors such as employer recommendations and a positive attitude, alongside negative predictors like being a health assistant, having poor knowledge, and working in a tertiary facility, provide a clear roadmap for intervention.

The findings underscore an urgent need for the implementation of robust, multi-level strategies, centered on mandatory institutional policies and targeted educational campaigns, to ensure the comprehensive protection of Nigeria’s healthcare workforce against this preventable occupational hazard.

## Declarations

### Ethics approval and consent to participate

Ethical approval was obtained from the Sokoto State Ministry of Health Research Ethics Committee (Ref: SMH/1580/V.IV) and the Usmanu Danfodiyo University Teaching Hospital, Sokoto, Ethics Committee (Ref: NHREC/30/012/2019). Written informed consent was obtained from all participants prior to their inclusion in the study. All methods were carried out in accordance with relevant guidelines and regulations.

### Consent for publication

Not applicable.

### Availability of data and materials

All data produced in the study are available upon reasonable request to the corresponding author.

## Data Availability

All data produced in the study are available upon reasonable request to the corresponding author.

